# Development and evaluation of patient-centred polygenic risk score reports for glaucoma screening

**DOI:** 10.1101/2024.09.25.24314343

**Authors:** Georgina L Hollitt, Mark M Hassall, Owen M Siggs, Jamie E Craig, Emmanuelle Souzeau

## Abstract

**Background:** Polygenic risk scores (PRS), which provide an individual probabilistic estimate of genetic susceptibility to develop a disease, have shown effective risk stratification for glaucoma onset. However, there is limited best practice evidence for reporting PRS and patient-friendly reports for communicating effectively PRS are lacking. Here we developed patient-centred PRS reports for glaucoma screening based on the literature and evaluated them with participants using a qualitative research approach.

**Methods:** We first reviewed existing PRS reports and literature on probabilistic risk communication. This informed the development of a draft glaucoma screening PRS report for a hypothetical high risk individual from the general population. We designed three versions of the report to illustrate risk using a pictograph, a pie chart and a bell curve. We then conducted semi-structured interviews to assess preference of visual risk communication aids, understanding of risk, content, format and structure of the reports. Participants were invited from an existing study, which aims to evaluate the clinical validity of glaucoma among individuals >50 years from the general population. Numeracy and literacy levels were assessed.

**Results:** We interviewed 12 individuals (50% female, 42% university education). Numeracy (mean 2.1±0.9, range 0-3), graph literacy (mean 2.8±0.8, range 0-4) and genetic literacy (mean 24.2±6.2, range -20-+46) showed a range of levels. We analysed the reports under three main themes: visual preferences, understanding risk and reports formatting. The visual component was deemed important to understanding risk, with the pictograph being the preferred visual risk representation, followed by the pie chart and the bell curve. Participants expressed preference for absolute risk in understanding risk, along with the written content explaining the results. The importance of follow-up recommendations and time to glaucoma onset were highlighted. Participants expressed varied opinions in the level of information and the colours used, which informed revisions of the report.

**Conclusions:** Our study revealed preferences for reporting PRS information in the context of glaucoma screening, to support the development of clinical PRS reporting. Further research is needed to assess PRS communication in other contexts and with other target audiences (e.g. referring clinicians), and its potential psychosocial impact in the wider community.

## INTRODUCTION

Polygenic risk scores (PRS) for glaucoma have shown effective stratification of disease risk and severity in recent years.^1-4^. PRS combine common variants associated with a trait or phenotype weighted by their effect size to generate an individual risk score. It reflects a probabilistic estimate of genetic susceptibility of an individual to develop a disease, and is a stratification, rather than a diagnostic tool. Clinical applications of glaucoma PRS include population-based or targeted screening for risk-stratified surveillance programs, improved triaging of glaucoma suspects, and personalised interventions ^5^. Despite growing research of the clinical utility of a glaucoma PRS, there is a lack of evidence of effective communication tools which will facilitate its implementation into clinical practice.

Effective risk communication is essential to improve patient understanding and recall of results, and lead to favourable health behaviours.^6-8^ Although genetic testing reports traditionally focus on monogenic variants, the communication of probabilistic information in the field of medicine generally, or in clinical genetics and genetic counselling specifically is not novel (e.g. non-invasive prenatal testing, recurrence risk for complex diseases such as psychiatric disorders)^9,10^. However fundamental differences in polygenic risk testing require careful consideration of risk communication (e.g. limitations related to the testing technology and analysis pipelines such as variants quality control, population reference and ancestry).

There is currently limited evidence for best practice reporting and communicating PRS and standardised guidelines are lacking. The Human Genetics Society of Australasia and The National Society of Genetic Counsellors have emphasised the importance of appropriate risk contextualisation and tailoring communication to individuals’ preferences.^11,12^ Careful consideration must also be given to the range of education levels, as well as health literacy and numeracy levels.^13,14^ Adequate communication of PRS results is critical for the clinical implementation of polygenic testing and promotion of consumer engagement. Here we developed patient-centred PRS reports for glaucoma screening based on the literature and using a qualitative research approach based on participants’ feedback.

## METHODS

### Review of existing PRS reports

There was limited literature on the most effective methods of reporting PRS and the perspectives of patients to different report styles. Risk stratification reporting using PRS has been mainly restricted to research or direct-to-consumer (DTC) settings. We first performed a review of existing PRS reports to assess design and the range of information provided. This included consideration of risk information, visual aids used to convey risk, colour formats and information included. PRS reports available in academic and commercial settings were identified by GLH and ES through PubMed, DTC company websites and internet search, and their content is summarised in Additional file 1. PRS reports covered a range of conditions but were mainly limited to inherited cancers, cardiovascular diseases and type 2 diabetes. Some reports were paper-based while others were web-based, including some that included interactive features to adjust risk based on lifestyle risk factors. Most categorised risk into groups (e.g. low, high) and some included integrated risk scores with other clinical risk factors. Risk was reported either as relative or absolute risk (5y, 10y or remaining lifetime), but all reports presenting relative risk also included absolute risk. All included some visual aid to present risk, although there was quite some variability in the visual representation of risk: the most common visual tools used were icon arrays and bell curves, while few included thermometers, scale bars, bar graphs or line graphs. Reports used a range of colours to differentiate risk, most often on a green (low risk)/yellow or grey (average)/red (high) scale, with other colours including pink for breast cancer risk, and blue or orange for patient’s risk.

Although standardised approaches for reporting PRS results are lacking, there is a breadth of literature on genetic and probabilistic risk communication. A summary of the key points highlighted by previous studies is presented in Box 1. Preferences for risk communication vary between individuals and risk perception is impacted by levels of numeracy and health literacy.^13,15-17^ Text and graphics should be kept simple and presume low levels of numeracy and health literacy.^16,17^ An overall consensus is to report risk in different formats, including graphical and written (verbal and numerical) to account for individual preferences and different literacy/numeracy levels.^16-24^ Individuals with lower numeracy tend to prefer visual aids and verbal labels (e.g. low/high risk) whereas those with higher numeracy favour numerical formats.^13^ When communicating numerical information, studies have reported improved comprehension when using frequencies, percentages or have found no difference.^13,15,21,24^ Absolute risk is usually preferred over relative risk which tend to exaggerate the perception of difference, especially when the absolute risk is small.^16-18,24^ However, absolute risk can lack comparison if not provided in the context of baseline risk.^20,24^ Finally, visual aids improve comprehension, however preferences for visual risk representation vary between studies: most preferred options are icon arrays or pictographs^16,19,22,24-26^, bar graphs,^18,22,24,25,27^ and pie charts.^22^ These recommendations align with a study from Brockman et al. who reviewed nine published PRS reports and emphasised the importance of visual elements, colours and test descriptions to interpret PRS of risk, and additional resources to account for different individuals’ levels of understanding.^28^

#### Box 1

**Key points for genetic risk communication**

- Use simple and plain language
- Keep information simple to emphasise key messages
- Present information in multiple formats (repetition)
  ∘ Graphical and written
  ∘ Numerical and verbal for written information
- Be consistent with numerical information
  ∘ Use of different format (e.g. percentage vs frequency)
  ∘ Use consistent denominators when reporting frequencies
- Absolute risk is preferred over relative risk
  ∘ Present absolute risk in the context of relative risk
  ∘ Present baseline risk
- Use visual display
  ∘ Icon array with small numerators and bar graph with large numerators

### Development of draft PRS reports

We developed three versions of a two-page glaucoma PRS report targeted to general population risk screening, with design and content informed by the literature (Box 1). Additional file 2 presents the three different versions, with the final page included in all three different 2-page versions. Each report contained the same information about the participant, their PRS, result explanation, test limitations, and information about PRS and glaucoma including symptoms, risk factors, treatment options and resources (Page 4 of Additional file 2). The report was developed to minimise technical medical/scientific language, while maximise understanding for people with low health literacy and numeracy. We provided results for a high hypothetical risk group (95^th^ polygenic risk percentile) and included recommendations for glaucoma eye screening 6 monthly. This was based on Australian national recommendations for individuals at risk of developing glaucoma to have regular eye checks,^29^ with frequency established based on our existing protocols.^1^

The three mock reports developed presented the same risk result using three different visual aids. Based on the literature, we chose (i) a bell curve to depict a relative risk, or (ii) an icon array or (iii) a pie chart to convey a participant’s absolute risk compared to the average population. The icon array was selected as it was one of the preferred options from the literature. We chose a pie chart over a bar graph as graph literacy can impact the level of understanding of bar graphs,^30^ while a pie chart is a format most individuals can understand.^22^ We chose to use blue to illustrate risk on the icon arrays and a blue (low risk)/orange (high risk) combination for the pie chart and bell curve to avoid confusion in individuals with red-green colour blindness, the most common form of colour blindness. A blue bar was positioned on the bell curve to represent population risk and a thicker red bar to represent the 95^th^ percentile of the general population. In addition to presenting risk graphically, we presented risk both as verbal (e.g. “Your result – high risk”) and numerical information. The numerical format provided a description of the risk both as a percentage (e.g. “Your result indicates your risk of developing glaucoma is approximately 7%”) and a frequency (e.g. “7 out of 100 people as you will develop glaucoma”). Finally we presented both the “patient’s risk” and the average population risk.

### Study sample and design

This study was approved by the Southern Adelaide Clinical Human Research Ethics Committee (SAC HREC, ethics approval 2020/HRE00968) and adhered to the Revised Declaration of Helsinki. Participants in the GRADE study (Genetic Risk Assessment of Degenerative Eye Disease) were invited via mail or email.^31^ GRADE is a prospective study that aimed to recruit 1,000 participants over the age of 50 years from the general population to evaluate the clinical validity of PRS testing for glaucoma in stratifying high and low risk individuals. Convenience and purposive sampling was used to ensure the involvement of various age groups within GRADE, as well as gender, education level, numeracy, and literacy. For this study, a target of 10-14 participants was set, subject to data saturation and the responses provided. All participants provided informed signed consent.

Participants were sent a short survey to complete online to collect demographic information (age, gender, education, ethnicity, colour blindness) and assess numeracy, graph and genetic literacy using validated tests. Numeracy was assessed using the Objective Numeracy Scale,^32^ graph literacy using the Short Graph Literacy Scale^30^ and genetic literacy using both the Genetic Literacy Fast Test^33^ and eight true/false statements based on existing measures and adapted to glaucoma (Additional file 3A). Finally, participants were asked what information they would like to see included in a report for glaucoma genetic risk with eight options and an open question for additional comments (Additional file 3B). Once the online survey was completed, participants were sent the three mock reports developed to have time to review them before being invited to do an online interview.

### Interviews and analysis

One-on-one semi-structured interviews were performed via telephone. The interview guide was developed and modified from existing literature (Additional file 4).^28^ The interviews were structured to cover five main themes: preference of visual risk communication aids, understanding of risk, influence of reports on risk perception and behaviour, usefulness of report content, and general report format and layout. Participants were first asked about their previous experience with receiving genetic reports or medical results, and any aspects of reports they had received in the past they did or did not find useful. This gave an insight into their baseline experience. The remaining themes were discussed for each figure to allow for clearer comparison between formats and participant reflection on their preferences. The content, layout and structure of the remaining aspects of the report were then assessed. This included the balance of text and visual elements, font and colours used. In assessing the most preferred reports, participants were given the opportunity to provide feedback as to how it could be improved. Interviews were performed until saturation of ideas and feedback was reached. Interviews and analysis were conducted by GLH. GLH is a female doctor with clinical and research experience in glaucoma and genetics and training in conversational interviewing who previously consented participants into the GRADE study. Participants were informed that the interviewer was conducting the project as part of her PhD and explained the rationale for the study. Data was audio recorded and coded based using notes taken during interviews. Deductive content analysis (GLH) was consistent with the interview guide.^34^ Interviews were performed until thematic saturation was reached.

## RESULTS

Twelve interviews were performed between April and September 2022. The mean duration of the interviews was 25 minutes and 50 seconds. The characteristics of the cohort are shown in Table 1. Half of the participants were female, all were of European ethnicity, 42% had a university education and none reported colour blindness. Participants had a range of numeracy, genetic and graph literacy as shown in Table 2. No participant had received a genetic report previously. Most participants commented that routinely, test results would be sent to the requesting healthcare provider, or would be sent to another relevant practitioner, rather than to the patient themselves.

**Table 1:** Characteristics of the cohort.

| ID | Age | Gender | Ethnicity | Education Level | Preference |
| --- | --- | --- | --- | --- | --- |
| A | ≥70 | Male | European | University | 1. Pictograph<br>2. Bell curve<br>3. Pie chart |
| B | 60-69 | Male | European | Vocational training | 1. Pictograph<br>2. Pie chart<br>3. Bell curve |
| C | 60-69 | Female | European | Secondary School | 1. Pie chart<br>2. Pictograph<br>3. Bell curve |
| D | ≥70 | Female | European | University | 1. Bell curve<br>2. Pictograph<br>3. Pie chart |
| E | ≥70 | Male | European | Vocational training | 4. Pictograph<br>5. Pie chart<br>1. Bell curve |
| F | 60-69 | Female | European | University | 2. Pie chart |
|  |  |  |  |  | 3. Bell curve<br>4. Pictograph |
| G | ≥70 | Female | European | Vocational training | 6. Pictograph<br>7. Pie chart<br>1. Bell curve |
| H | 60-69 | Female | European | University | 2. Pie chart<br>3. Pictograph<br>4. Bell curve |
| I | 60-69 | Male | European | University | 1. Pie chart<br>2. Pictograph<br>3. Bell curve |
| J | 60-69 | Male | European | Secondary School | 1. Pictograph<br>2. Pie chart<br>3. Bell curve |
| K | 60-69 | Male | European | Vocational training | 1. Pictograph<br>2. Pie chart<br>3. Bell curve |
| L | 60-69 | Female | European | Secondary School | 1. Pictograph<br>2. Pie chart<br>3. Bell curve |

**Table 2:** Participants’ numeracy, graph and genetic literacy.

| Survey |  |
| --- | --- |
| **Numeracy score**<br>Mean (SD)<br>Range (0-3)<br>All answers correct | 2.1 (0.9)<br>0-3<br>4/12 (25.0%) |
| **Genetic literacy score**<br>Mean (SD)<br>Range (-20 to 46)<br>Low genetic literacy (≤23) | 24.2 (6.2)<br>14-39<br>6/12 (50%) |
| **Genetic knowledge**<br>Mean (SD)<br>Range (0-8)<br>All answers correct | 7.0 (1.0)<br>5-8<br>4/12 (33.3%) |
| **Graph literacy score**<br>Mean (SD)<br>Range (0-4)<br>All answers correct | 2.8 (0.8)<br>2-4<br>2/12 (16.7%) |

A summary of the participants’ preferences for the graph format reporting results is summarised in Table 3. The most popular first preference was the pictograph format, followed by the pie chart and lastly the bell curve. Most elements were deemed important by participants, apart from information about insurance which only a third of participants wished to see on the reports (Figure 1). In analysing preferences for the format of representing risk and information, three main themes were identified to contribute to overall understanding of the reports developed. Firstly, preferences towards the visual component used to visually represent the risk, which included the format of the figure and the presentation of risk as either absolute or relative. Secondly, accuracy of understanding and confidence in interpreting the visual component together with the accompanying text explaining the result. Thirdly, the informative text providing more detail about the test and glaucoma, together with the overall format and layout of the reports.

**Table 3:** Participants’ preferences for graph format.

| Graph format | First preference | Second preference | Third preference |
| --- | --- | --- | --- |
| Pictograph | 7 (58.3%) | 4 (33.3%) | 1 (8.3%) |
| Pie chart | 4 (33.3%) | 6 (50.0%) | 2 (16.7%) |
| Bell curve | 1 (8.3%) | 2 (16.7%) | 9 (75.0%) |

**Figure 1:**
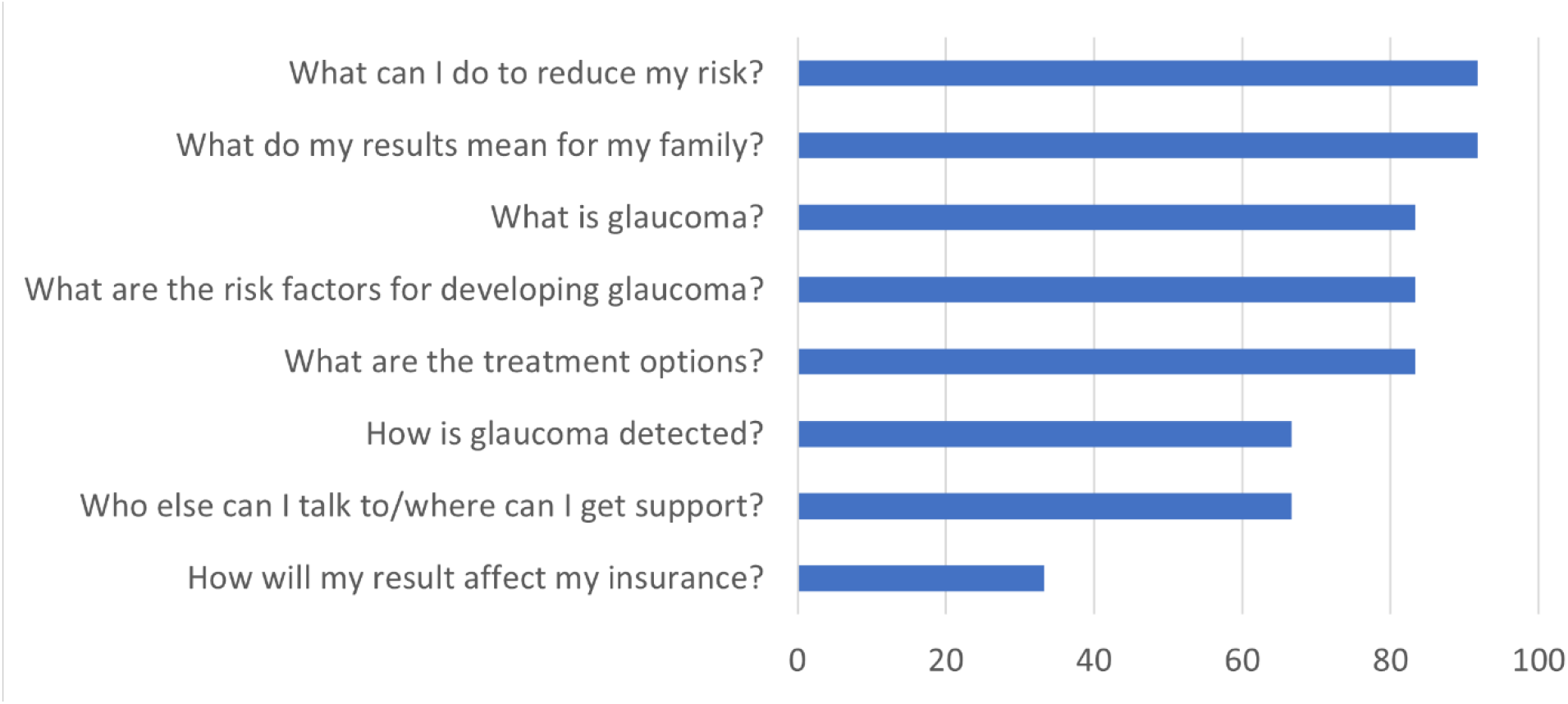
Proportion of participants who selected elements deemed important to include in the report. Participants could select more than one option.

### Theme One: Preferences towards visual risk communication aid

Overall, absolute risk was preferred, either in the format of pictographs or a pie chart, with the bell curve being the least preferred option. The two absolute risk figures helped participants understand their risk by visually comparing the personal risk to the general population.

Most participants felt the pictograph was visually clear and could be interpreted quickly, without needing the corresponding text to help understand the result.

> *‘Clear and simple’*
>
> *‘I don’t think you have to think about it, it’s [the result] there in front of you…it’s very definitive in its message’*

Additionally, participants were more readily able to correctly interpret the result.

> *‘That I’m more than twice as likely to get glaucoma than the general population’*
>
> *‘You can see that you’re at higher risk than the average (population), but you’re not 95% like the other one gives the impression of’*

Participants felt the pie chart was easy to interpret, mainly because of its clear comparison to the average population.

> *‘Very clear, you don’t have to think about it’*
>
> *‘I didn’t need the (corresponding) text as much’*

The bell curve was less effective in helping participants conceptualise their risk. Most commented that this figure gave the impression of extremely high risk, or almost certainty, of developing glaucoma and would therefore cause significant worry. The relative risk, presented as a percentile with the bell curve, was difficult to understand for some.

> *‘I don’t really get it. Don’t even go there’*
>
> *‘It’s hard to get it all in the head…and work it out’*
>
> *‘I don’t think [other people would understand]…you have to look at it’*

The bell curve was the preferred visual aid for one participant, who prefaced their feedback by noting having had quite a lot of experience with interpreting bell curves in the past and therefore being very comfortable with this format. However, this participant felt most people would not be able to understand this figure.

> *‘The picture matches the words underneath. I think it’s easy to understand’*
>
> *‘I think that a lot of people are probably not familiar with looking at themselves within a population’*

### Theme Two: Understanding risk

The visual component had a significant impact on understanding and assisting in translating visual risk to numerical risk. Overall, participants reported that the absolute risk figure would give more understanding without making the individual feel too worried. The bell curve was felt to be most confusing, and was generally misinterpreted as a percentage risk of developing glaucoma. Most participants did not fully understand the concept of a percentile to represent risk within a population, compared to a percentage.

> *‘I looked at the graph first and went, oh 95%, and then I read it…and realised it was 2*.*3 times higher, not 95% chance’*

Confusion and misinterpretation of the risk presented influenced the degree of worry and risk perception. Participants felt that the absolute risk represented by the pictograph and pie charts were reassuring, and while each figure reported the same high-risk result, these two figures represented a much lower risk compared to the bell curve. Some felt that this may negatively affect risk-reducing behaviour.

> *‘It still…can indicate, compared to the rest of the population, at a relatively low risk of developing glaucoma. Whether or not people would act on that’*

All participants felt the corresponding written content underneath the graph, explaining the result, was useful and necessary to aid understanding. Although participants often felt their understanding of the report was sound, many felt a recommended follow-up timeframe would significantly contribute to their behaviour by indicating urgency in terms of wanting to review the report with a healthcare professional, such as their optometrist or GP, undergoing an eye examination, or discussing their result with their family.

> *‘The main thing I want to know is what to do with my result. What do I need to do next and when?’*
>
> *‘I was happy with the content - people want to know what it means for them and where to go next*.*’*

### Theme Three: Report format and visual elements

The visual and design elements played an important role in facilitating understanding and risk perception. Particularly, they contributed most to a user’s first impression.

> *‘The first thing you look at is the visual, and then you read*.*’*

Colour was a predominantly discussed design element, which contributed to confusion for some participants with the bell curve. Participants felt that the blue-orange colour scheme did not make sense initially, and negatively influenced their overall understanding and experience with this graph.

> *‘The colours too…didn’t make a statement’*
>
> *‘I think that the shading probably makes it a little more confusion…the shading make it less definitive’*
>
> *‘I can see how you’ve faded the colours, gone from the caution colour to the cool colour, but I didn’t pick up on that immediately*.*’*

All participants felt the font used was appropriate and of adequate size, particularly given reports may be read by individuals with visual impairment. There was little feedback on this aspect.

Participants generally felt the layout of the report was simple and easy to follow. Bullet points were useful in communicating relevant information without too much detail, using simple language. Most participants felt there was an appropriate balance of text and visual elements. Most participants felt the content of the report was appropriate, however all wanted further detail and emphasis on follow-up or treatment recommendations. One participant felt there was too much information included.

> *‘The section I thought was over the top was those second and third sections, that’s a lot of text. People are just not going to read it*.*’*

A suggested modification to improve this was to include more detailed information as smaller text at the end of the report.

> *‘You could have, in a lot smaller print, on the back of the pamphlet the limitations of the test and all of those sorts of stuff that you need to perhaps tell people, but it’s not the primary objective of the result*.*’*

### Potential modifications based on feedback

Based on the feedback received from these interviews, a number of modifications to our reports could be made. Section three in the report draft could be moved to follow the reported PRS result, to further improve and support their understanding of the results. While the colour scheme of orange and blue was chosen to aid interpretation of those with red-green colour blindness, all participants felt another colour scheme would add to the visual interpretation. A red-green colour scheme was suggested, which is familiar to most people in settings such as traffic lights and temperature gradients. Improving understanding for a larger majority may be more useful in achieving greater understanding, although it would come at the sacrifice of the smaller number of those with colour blindness.

## DISCUSSION

Glaucoma PRS have many potential clinical applications,^5^ and are already being used in clinical practice. Previous research from our group showed positive attitude toward polygenic risk testing among affected and unaffected individuals, as well as support from healthcare professionals.^35-37^ As clinical implementation occurs, the ability to effectively and efficiently report and communicate results will be essential. Interpretation of results involves non-genetic healthcare providers (e.g. ophthalmologists, optometrists) and consumers/patients themselves, and as such requires scalable models.^6^ Previous research supports a user-centred approach to the design of genetic test reports for optimal comprehension and communication.^38-41^ Although there are currently no best practice guidelines to report PRS results specifically, professional genetic organisations have emphasised the importance of involving target groups in the co-design and evaluation of risk communication resources.^11,12^ In this qualitative study, we developed patient-centred PRS reports for glaucoma screening based on a review of the literature and surveyed participants’ preferences and feedback.

While there are some fundamental differences with monogenic testing, the communication of individualised risk is not new to medicine or genetics, and there exists a breadth of literature on genetic risk communication involving probabilities.^13,15-27^ In this study, we reviewed existing PRS reports for various conditions. There was significant variability in the design of the reports, however common themes were identified. Generally, graphic design was included to aid comprehension, relative risk was presented in the context of absolute risk, and risk groups were usually reported. Graphical displays are known to facilitate risk communication and promote healthy behaviours.^42,43^ Similarly, previous research has emphasised that absolute risk is usually preferred over relative risk^16-18,24^, risk should be contextualised in comparison to the general population,^20,24^ and a preference for simplified visual aids such as pictographs over graphical representations.^16,19,22,24-26^ Here, the participants validated the importance of the visual components for understanding results. Our findings highlighted similar preferences of pictographs and pie charts depicting absolute risk over a bell curve representing relative risk, while expressing the importance of providing baseline or population level risk to conceptualise the individual risk.

One of the most significant challenges to consider in designing PRS reports is the variation in literacy and numeracy levels within the target population. Public familiarity with genomic risk information is generally low.^44-46^ Moreover, risk understanding and health outcomes are known to be associated with levels of numeracy, graph, health and genetic literacy.^30,33,47,48^ Lower numeracy can lead to overestimation of risk and lower ability to use risk reduction information.^13^ We assessed numeracy, graph and genetic literacy levels in this study. Although the cohort had a high level of education compared to the Australian population,^49^ there was a range of numeracy and literacy levels.^32,33,50^ We developed reports that included simple graphics and text, and risk representation in graphical as well as written verbal and numerical formats, to account for low levels of numeracy and literacy. Participants overall reported difficulty understanding the bell curve representing a relative risk. Our findings support the concept that relative risk leads to higher risk perception in the context of a relatively low absolute risk.^16-18,24^ Further research is needed to assess how risk communication and perception might impact behavioural changes.

Results disclosure usually involves communicating the implications of the results. There are currently no clinical guidelines for the surveillance or management of glaucoma based on PRS results or risk groups. In this study, participants were provided with the current Australian recommendations for the general population to have regular eye health checks,^29^ with frequency of screening specified from our exiting protocols.^1^ Healthcare providers have previously expressed concern over the lack of clinical practice guidelines for the implementation of PRS in practice,^51^ including in the context of glaucoma.^37^ The importance of clear recommendations based on genetic risk was highlighted by participants in our study, especially for guiding an individual’s behaviour in response to their genetic risk. The development of clinical guidelines to provide appropriate advice to patients and their clinicians for different risk groups has been identified as a priority by professional genetic organisations,^11,12^ and is currently being addressed by prospective studies.

Strengths of this study include the development of PRS reports based on a review of the literature of risk communication, and existing PRS reports, with potential improvements informed by feedback from participants with a range of numeracy and literacy. Limitations include a cohort that was highly educated and of European background, which may not represent the target population for PRS-based glaucoma screening. We also did not survey individuals from other geographical, cultural, and linguistic groups, nor did we develop or test reports for other clinical contexts (e.g. glaucoma cases, family members), or other target audiences (e.g. referring clinicians). Finally, the study assessed a mock report representing a hypothetical high-risk result, and outcomes may differ for low or intermediate results. In addition, a real personalised risk result might be perceived differently by individuals.

## CONCLUSIONS

This study provided a framework for the disclosure of PRS results in the context of glaucoma screening with a patient-centred report. Further research should assess PRS communication in a broader range of target populations and clinical contexts, including the potential psychosocial impact of returning personalised risk using such reports.

## Data Availability

Data generated or analysed during this study are included in this published article and its supplementary information files.

---

**Additional File 1:** Review of existing polygenic risk score reports formatting and content

**Additional File 2:** Polygenic risk score reports

**Additional File 3:** Participants literacy questionnaire (A) and preferences for reports’ content (B)

**Additional File 4:** Interview semi-structured interview guide

