## Additional File 1 for "Development and evaluation of patient-centred polygenic risk score reports for glaucoma screening"

**Additional file 1:** Review of polygenic risk score reports formatting and content

| Study/Provider | Condition(s) reported/selected | Presentation of risk (inc. absolute vs relative, percentile vs risk groups) | Visual aids (presence, format, colour) | Information included | Reference |
|---|---|---|---|---|---|
| Fenton et al. | ▪ Melanoma | ▪ Absolute risk (remaining lifetime)<br>▪ Relative risk (compared to others same age & sex)<br>▪ Risk group (low, average, high) | ▪ Icon array (100 person) | | 1 |
| Forrest et al. | ▪ Breast cancer | ▪ Relative risk<br>▪ Absolute risk (lifetime)<br>▪ Risk group (ex: high) | ▪ Compared 2 visual tools<br>▪ Bell curve showing patient (red) & population risk (green), lifestyle risk factors (alcohol & obesity) & BRCA risk<br>▪ Icon array of 100 women (pink, full symbols for patient's risk) | ▪ Methods and genomic variants tested | 2 |
| Brockman et al. | ▪ Coronary artery disease | ▪ Relative risk (inc. percentile)<br>▪ Risk group (reduced, average, increased)<br>▪ Population risk | ▪ Bell curve showing patient's risk in green (low), grey (average) or red (high) | ▪ Information about condition, family testing, recommendations, PRS calculation, insurance implications | 3 |
| P5 Study | ▪ Type 2 diabetes, coronary heart disease, venous thrombosis | ▪ Absolute risk (10y)<br>▪ Risk group (low, increased, high, very high)<br>▪ Risk calculator (inc. lifestyle risk factors) | ▪ Thermometer & bell curve with person diagram (green/yellow/pink)<br>▪ Bar graph showing patient vs population risk at current age & patient's risk in 10y | ▪ Information about condition, lifestyle recommendations, test methodology | 4 |
| CanRisk | ▪ Breast & ovarian cancer | ▪ Absolute risk (5y, 10y & remaining lifetime)<br>▪ Risk group (near population, moderate, high) | ▪ Icon array<br>▪ Line graph showing patient vs population risk (blue/grey) | | 5 |
| GeneRISK Study | ▪ Atherosclerotic cardiovascular disease | ▪ Absolute risk (10y, lifetime)<br>▪ Risk group (low, moderate, high) | ▪ Scale bar showing patient's risk (orange)<br>▪ Line graph showing patient's risk (orange) compared to average population (blue) | ▪ Modifiable risk factors | 6 |
| eMERGE study | ▪ Breast cancer<br>▪ Type 2 diabetes (selected) | ▪ Relative risk<br>▪ Absolute risk (lifetime)<br>▪ Risk group 9high, not high) | ▪ Icon array<br>▪ Bell curve showing patient's risk (red) | ▪ Information about condition, recommendations, test methodology & limitations | 7 |

| | | | | | |
|---|---|---|---|---|---|
| 23andMe | ▪ Coronary artery disease (selected) | ▪ Absolute risk (by certain age)<br>▪ Risk group (ex: increased) | ▪ Scale bar (green/yellow/red) | ▪ Information about condition, lifestyle recommendations, test methodology & limitations (ancestry) | 8 |
| MyGeneRank | ▪ Coronary artery disease (example) | ▪ Absolute risk<br>▪ Risk group (ex: moderate) | ▪ Scale bar (blue/red) | ▪ Recommendations | 9 |
| Myriad Genetics | ▪ Breast cancer (example) | ▪ Absolute risk (remaining lifetime, compared to population) | ▪ Bar graph showing patient vs population risk (pink/grey) | ▪ Test methodology, recommendations | 10 |
| Ambry Genetics | ▪ Breast cancer<br>▪ Prostate cancer | ▪ Relative risk<br>▪ Absolute risk (lifetime, compared to population) | ▪ Bar graph comparing patient (pink/blue) to baseline (grey) | ▪ Test methodology, recommendations left to the discretion of healthcare provider | no longer available |
