## Additional File 2 for "Development and evaluation of patient-centred polygenic risk score reports for glaucoma screening"

### Polygenic Risk Score (PRS) Report - Glaucoma

ID: xxxxxxxxxxx

First Name: ……………………………………………………
Surname: ……………………………………………………
DOB: ……………………………………………………
Gender: ……………………………………………………

Ordered by: ……………………………………………
Sample collection date: ……………………………
Received at lab: ……………………………………
Date of reporting: ……………………………………

#### 1. PRS Result - Glaucoma

##### Your result - high risk

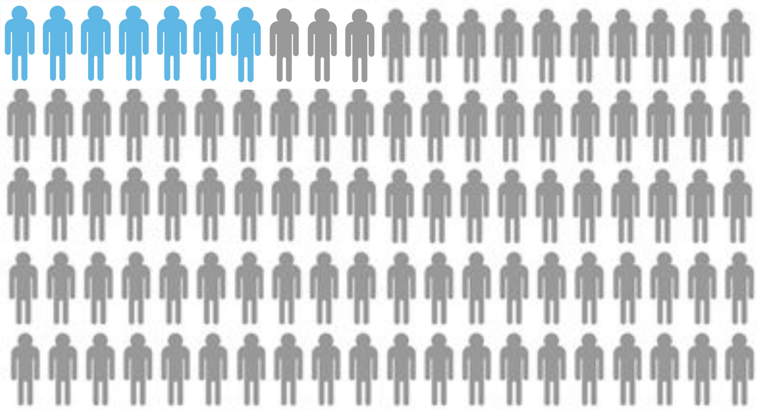

Your result indicates your risk of developing glaucoma is approximately 7%. This means that 7 out of 100 people of the same gender and age as you will develop glaucoma over their lifetime.

##### Average population

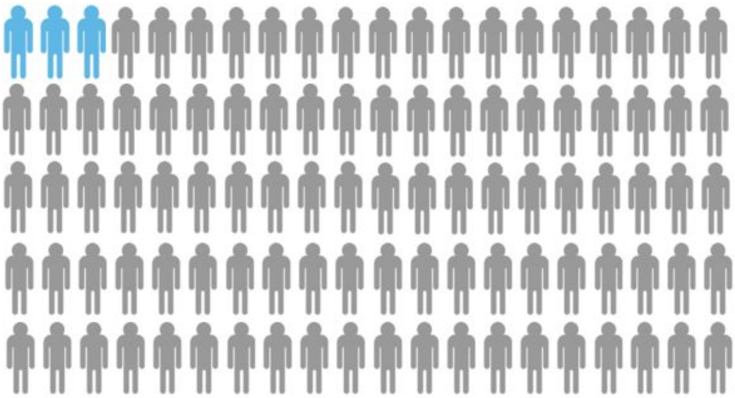

Average population - the average population risk is approximately 3%. This means that on average, 3 in 100 people of the same age and gender as you will develop glaucoma.

#### 2. Polygenic Risk Scores (PRS) explained

- Polygenic risk scores give an indication of a person's overall genetic risk for a particular condition.
- Your test results were based on 2673 genetic changes (or variants) that we know influence a person's risk of developing glaucoma.
- Some variants are more strongly associated with glaucoma than others.
- Polygenic risk scores add up all of the genetic variants associated with a particular condition that a person has, accounting for how strongly they are associated with the condition.
- Your score (and genetic risk) can be compared to other people within the population.

**What are the limitations of the test?**

- This test estimates your risk of primary-open angle glaucoma (the most common subtype of glaucoma).
- It does not estimate your risk of other types of glaucoma or other conditions.
- Although the polygenic score predicts risk in many ancestries, it has been best validated in individuals of European ancestry.
- This test does not account for some rare variants known to cause glaucoma. Therefore, your risk may be higher, especially if you have a strong family history of glaucoma.
- PRS results represent a probability of individual disease risk and are therefore not diagnostic, and results should be interpreted by a suitably qualified clinician in conjunction with other established clinical risk factors, in particular age. It does not take into account non-genetic risk factors.

#### 3. What does my test result mean for me?

- Your result suggests you are at 2.3 times higher risk of developing glaucoma than most other people.
- Your result does not mean that you have glaucoma now.
- It does not mean you will definitely develop glaucoma.
- Because you are at increased risk, we recommend that you see an *ophthalmologist* or *optometrist* every *6 months* so they can check your eyes to see if you have glaucoma.
- There are several treatment options available for glaucoma. Please refer to the back of the page for more information on glaucoma and treatment options.
- Your result also means that other people in your family may be at increased risk as well. They can talk to their GP or eye care professional about it.

### Polygenic Risk Score (PRS) Report - Glaucoma

ID: xxxxxxxxxxx

First Name: ………………………………………………………
Surname: …………………………………………………………
DOB: ………………………………………………………………
Gender: ……………………………………………………………

Ordered by: ……………………………………………………
Sample collection date: ……………………………………
Received at lab: ………………………………………………
Date of reporting: ……………………………………………

#### 1. PRS Result - Glaucoma

**Your result - high risk**

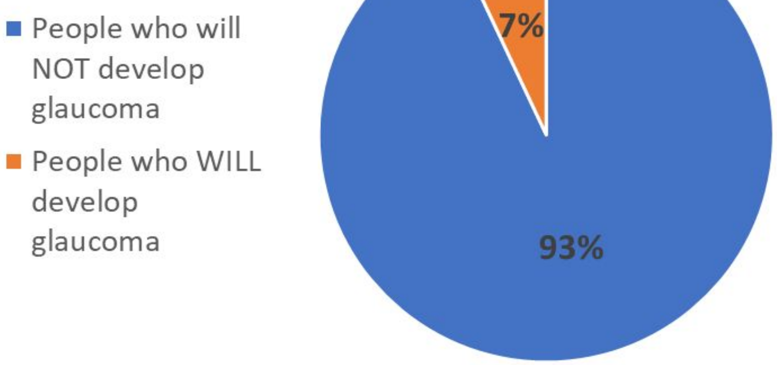


**Average population**

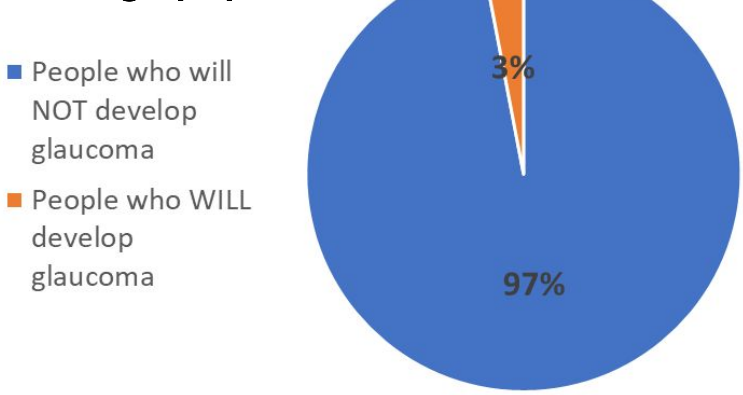


Your result indicates your risk of developing glaucoma is approximately 7%. This means that 7 out of 100 people of the same gender and age as you will develop glaucoma over their lifetime.

Average population - the average population risk is approximately 3%. This means that on average, 3 in 100 people of the same age and gender as you will develop glaucoma.

#### 2. Polygenic Risk Scores (PRS) explained

- Polygenic risk scores give an indication of a person's overall genetic risk for a particular condition.
- Your test results were based on 2673 genetic changes (or variants) that we know influence a person's risk of developing glaucoma.
- Some variants are more strongly associated with glaucoma than others.
- Polygenic risk scores add up all of the genetic variants associated with a particular condition that a person has, accounting for how strongly they are associated with the condition.
- The score is represents your risk compared to other people within the population.

**What are the limitations of the test?**

- This test estimates your risk of primary-open angle glaucoma (the most common subtype of glaucoma).
- It does not estimate your risk of other types of glaucoma or other conditions.
- Although the polygenic score predicts risk in all ancestries, it has been best validated in individuals of European ancestry.
- This test does not account for some rare variants known to cause glaucoma. Therefore, your risk may be higher, especially if you have a strong family history of glaucoma.
- PRS results represent a probability of individual disease risk and are therefore not diagnostic, and results should be interpreted by a suitably qualified clinician in conjunction with other established clinical risk factors, in particular age. It does not take into account non-genetic risk factors.

#### 3. What does my test result mean for me?

- Your result suggests you are at 2.3 times higher risk of developing glaucoma than most other people.
- Your result does not mean that you have glaucoma now.
- It does not mean you will definitely develop glaucoma.
- Because you are at increased risk, we recommend that you see an *ophthalmologist* or *optometrist* every *6 months* so they can check your eyes to see if you have glaucoma.
- There are several treatment options available for glaucoma. Please refer to the back of the page for more information on glaucoma and treatment options.
- Your results also means that other people in your family may be at increased risk as well. They can talk to their GP or eye specialist about it.

### Polygenic Risk Score (PRS) Report - Glaucoma

ID: xxxxxxxxxxx

First Name: ………………………………………………………
Surname: ………………………………………………………
DOB: ………………………………………………………
Gender: ………………………………………………………
Ordered by: ………………………………………………
Sample collection date: ……………………………………
Received at lab: ……………………………………………
Date of reporting: …………………………………………

#### 1. PRS Result - Glaucoma

**Your result - high risk**

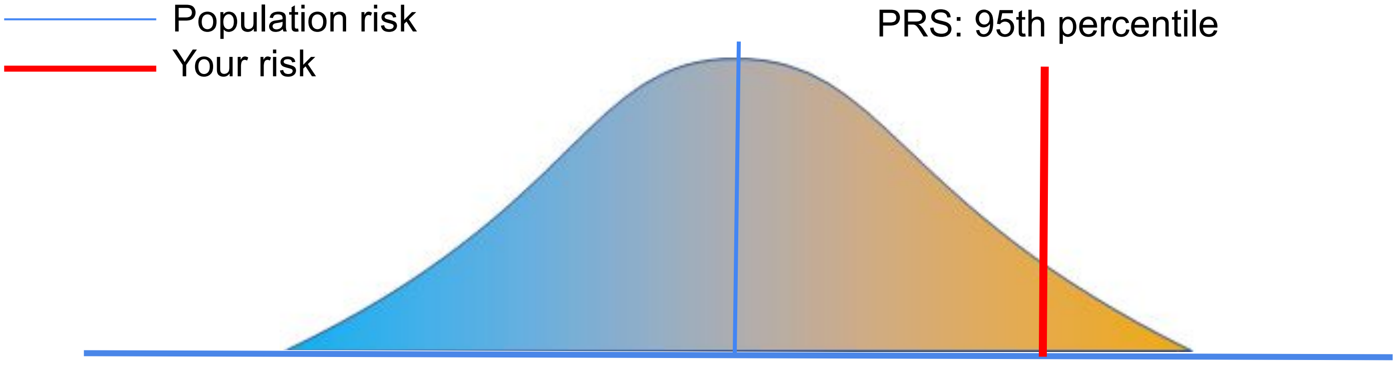


Your Glaucoma PRS indicates you are in the 95th percentile of risk within the population. This means that your risk of developing glaucoma is higher than 95% of the population. Out of 100 people, you have a higher genetic risk of glaucoma than 95 people.

#### 2. Polygenic Risk Scores (PRS) explained

- Polygenic risk scores give an indication of a person's overall genetic risk for a particular condition.
- Your test results were based on 2673 genetic changes (or variants) that we know influence a person's risk of developing glaucoma.
- Some variants are more strongly associated with glaucoma than others.
- Polygenic risk scores add up all of the genetic variants associated with a particular condition that a person has, accounting for how strongly they are associated with the condition.
- The score is represents your risk compared to other people within the population.

**What are the limitations of the test?**

- This test estimates your risk of primary-open angle glaucoma (the most common subtype of glaucoma).
- It does not estimate your risk of other types of glaucoma or other conditions.
- Although the polygenic score predicts risk in all ancestries, it has been best validated in individuals of European ancestry.
- This test does not account for some rare variants known to cause glaucoma. Therefore, your risk may be higher, especially if you have a strong family history of glaucoma.
- PRS results represent a probability of individual disease risk and are therefore not diagnostic, and results should be interpreted by a suitably qualified clinician in conjunction with other established clinical risk factors, in particular age. It does not take into account non-genetic risk factors.

#### 3. What does my test result mean for me?

- Your result suggests you are at 2.3 times higher risk of developing glaucoma than most other people.
- Your result does not mean that you have glaucoma now.
- It does not mean you will definitely develop glaucoma.
- Because you are at increased risk, we recommend that you see an *ophthalmologist* or *optometrist* every *6 months* so they can check your eyes to see if you have glaucoma.
- There are several treatment options available for glaucoma. Please refer to the back of the page for more information on glaucoma and treatment options.
- Your results also means that other people in your family may be at increased risk as well. They can talk to their GP or eye specialist about it.

### Polygenic Risk Score Report - Glaucoma

ID: xxxxxxxxxxx

#### Your Polygenic Risk in Detail

- Researchers have identified genetic variants which are associated with glaucoma by comparing those with the disease to those without
- A PRS collates the combined risk of multiple genetic risk variants into a single score, typically by weighting the relative effect size of each variant.
- Scores may be combined with conventional risk factors to estimate overall disease risk.

For a full explanation on genetic risk and calculation of PRS:
https://www.genome.gov/Health/Genomics-and-Medicine/Polygenic-risk-scores

#### Frequently Asked Questions

**What is glaucoma?**

- Glaucoma is a group of neurodegenerative conditions that affect the optic nerve
- Glaucoma is usually a complex disease, influenced by both genetic and environmental factors.
- Primary open-angle glaucoma is the most common form of glaucoma
    - Open-angle means the area where the fluid drains out of the eye is
    - not obstructed
    - Primary means there is no other known cause (such as trauma or surgery)

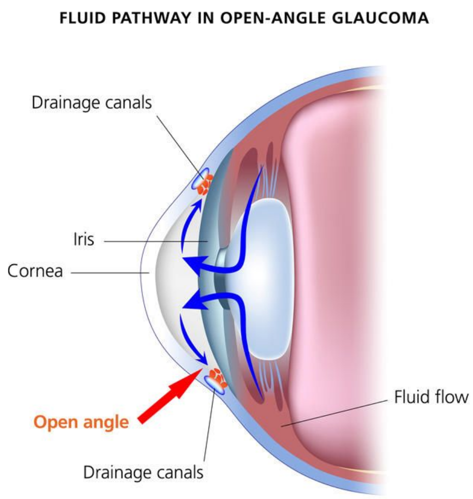


**What are the symptoms?**

- Usually there are no symptoms in early disease
- Vision loss may only be noticeable in later stages of disease
- Vision loss from glaucoma is irreversible and cannot be restored
- Only an eye exam performed by an eye specialist can tell if someone has glaucoma

**Are there any risk factors?**

- Yes, there are a number of risk factors (examples listed below)
- Family history of glaucoma
    - Those who have a first-degree relative (parent, sibling, child) with glaucoma are at almost 10 times increased risk of also developing glaucoma compared to those who do not
- African ancestry
- Age over 50 years
- Elevated eye pressure

**What are the treatment options?**

- Treatment options are highly effective at slowing or preventing disease progression in most people
- Treatments include topical eye drops, laser therapy, and in very advanced cases, surgery.

**What does this mean for my family?**

- We know that people who have a family member with glaucoma are at higher risk of also developing glaucoma
- If you have been identified to be at high risk, it is possible that your closest relatives are also at increased risk

#### Resources - for more information and where to get help

Glaucoma Australia:
https://glaucoma.org.au/home

Glaucoma Australia
Saving sight

Vision Australia:
https://www.vision2020australia.org.au/

VISION 2020
THE RIGHT TO SIGHT
AUSTRALIA

Speak to your:
- Ophthalmologist
- Optometrist
- GP
