## Additional File 3 for "Development and evaluation of patient-centred polygenic risk score reports for glaucoma screening"

**Additional file 3:** Participants literacy questionnaire and preferences for reports' content

A. Knowledge of glaucoma risk. Participants were asked to answer 8 true/false statement to assess their health literacy.

Indicate whether you think the following statements are either 'true' or 'false'.

1. Most genetic disorders are caused by a single gene
2. A "complex disease" is a health condition brought on by many genes and lifestyle and environment
3. If your close relatives have glaucoma, you are more likely to develop it
4. Glaucoma screening is only recommended for people with a family history of glaucoma
5. Each of us has variations in our genes that make it more likely that we will get certain diseases
6. If a person has a genetic predisposition for a disease, this person will always get the disorder
7. The exact chance of developing a genetic condition can be determined through genetic testing
8. Once a genetic marker for a disorder is identified in a person, the disorder can usually be prevented or cured

B. Participants' content preferences. Participants were asked to select information they would like to see included in the reports.

In preparing for discussing the reports, could you tell us which of the following questions and explanations would you want to see included in a report for glaucoma genetic risk? (choose as many as appropriate)

1. What is glaucoma?
2. What are the risk factors for developing glaucoma?
3. What can I do to reduce my risk?
4. How is glaucoma detected?
5. What are the treatment options?
6. What do my results mean for my family?
7. Who else can I talk to/where can I get support?
8. How will my result affect my insurance?

Other (please specify)
