## Additional File 4 for "Development and evaluation of patient-centred polygenic risk score reports for glaucoma screening"

**Additional file 4:** Interview semi-structured interview guide

<u>Discussion points:</u>

- Overall impression of the different reports
- Impact of colour on report preference
- Impact of font on report preference
- Whether the report content is appropriate
- Whether the report content is easy to understand
- Whether there was anything missing from the reports
- Does the report raise any questions
- What do they understand about the representation of the risk
- Whether the report adequately communicates the results to ensure the individual would feel confident that they have correctly understood it.

**Introduction and Purpose**

Thank you for volunteering and taking the time to participate in this project. My name is Georgie, and I am conducting this study as part of my PhD within the Department of Ophthalmology at Flinders University. You have been asked to participate as your view is important. We appreciate your time and willingness to participate. This interview is designed to explore your thoughts and preferences towards 3 reports which we have recently created to communicate polygenic risk score results to members of the community.

As you may remember, the GRADE study is working towards integrating polygenic risk testing for glaucoma into clinical practice, so it will be important to ensure that the results are reported effectively. That is why we are asking for your honest opinions.

There are absolutely no right or wrong answers. We want to hear any feedback you may have.

We are recording your answers so that we don't miss anything, but your answers will be kept confidential. The recordings are stored safely. When they are transcribed, nobody's name will be attached to their comments.

Can I ask you again for your permission to record the discussion? *[Upon yes, start recording]*

**General**

- Have you ever received a genetic report before? If yes, did you find it useful?
- Have you ever received any other medical results in a written report before?
    - If yes, did you find it useful?
    - What did you like about the report?
    - What didn't you like about the report

**First Impressions**

- What is your first impression of the report?
- What is the first thing you looked at on the report?

- Is there anything you would suggest changing to improve your first impression of the report?
- After reading the report, can you tell me what the main thing this report is telling you?

**Risk figure (each):**

- Do you think this figure is easy to understand? If not, what is difficult to understand?
- What do you like about this figure?
- What don't you like about this figure?
- What is the key message of the figure?
- What is your interpretation of the risk that is shown in the figure? Do you think this person is at higher risk, lower risk, or average risk of developing glaucoma compared to other people?
- On a scale of 1-10, How worried would you be if you saw this result, with 1 being not at all worried and 10 being extremely worried?
- Do you think most people would be able to understand this figure? Why/why not?

**Preference:**

- Out of the three figures you have been shown, which did you prefer? Why?
- Can you rank the three figures in order of most preferred to least preferred?
- Can you rank the three figures in order of easiest to hardest to understand?
- Do you have any suggestions about how they could be improved?
- Do you have any further comments about these figures?

**Report layout and appearance**

- Do you think the report was easy to read and interpret?
- Do you think the content of the report was appropriate?
- Was there any information missing from the reports that you would like to see?
- What did you like about the report?
- What did you not like about the report?
- Can you comment on the balance of text and visual elements? Is there too much of one and not the other?
- Did you like the font that was used?
- Did you like the colours that were used?

**Confidence**

- Would you feel confident that you correctly understood the report?
- Would you want to review the report with your doctor?
- Do you have any other comments/feedback on the report/s?

**Summary**

Thank you very much for sharing your thoughts and preferences on the reports. We are reaching the end of our time now, so I would like to finish by summarising the key ideas that I have heard.

Is there anything I have missed or anything anyone/you would like to add?

If you think of anything later that you would like to feed back, you are welcome to contact me via email or phone. My contact details can be found on the Information Sheet.

Thank you again for your contribution to this project. Your honest discussion has been very helpful in furthering our understanding of how we can effectively communicate genetic results for glaucoma to members of the community.

**Useful prompts to use throughout interview:**

- Can you tell us a little bit more about that?
- Can you give an example of what you mean?
- For negative responses
- Can you give a suggestion on how it could be changed/improved?
